# Projections of human papillomavirus (HPV) vaccination impact in Ethiopia, India, Nigeria, and Pakistan: a comparative modelling study

**DOI:** 10.1101/2021.06.01.21258170

**Authors:** Allison Portnoy, Kaja Abbas, Steven Sweet, Jane J. Kim, Mark Jit

## Abstract

**Background:** Cervical cancer is the second most common cancer among women in Ethiopia, India, Nigeria, and Pakistan. However, of these four countries, only Ethiopia has introduced human papillomavirus (HPV) vaccination at the national level in 2018 and India in a few states in 2016. Our study objective was to estimate the potential health impact of HPV vaccination among ten cohorts of 9-year-old girls from 2021–2030 in Ethiopia, India, Nigeria, and Pakistan using two independent mathematical models, and assess similarities and differences in vaccine impact projections through comparative modelling analysis.

**Methods:** Using two widely published models (Harvard and PRIME) to estimate HPV vaccination impact, we simulated a vaccination scenario of 90% annual coverage among 9-year-old girls from 2021–2030 in Ethiopia, India, Nigeria, and Pakistan. We estimated the potential health impact in terms of cervical cancer cases, deaths, and disability-adjusted life years (DALYs) averted among vaccinated cohorts from the time of vaccination until 2100. We also conducted a comparative modelling analysis to understand the differences in vaccine impact estimates generated by the two models.

**Results:** Prior to harmonising model assumptions, the range between the PRIME model and the Harvard model for the potential health impact of HPV vaccination in terms of the number of cervical cancer cases averted among girls vaccinated 2021–2030 between the year of vaccination and 2100 was: 262,000 to 270,000 in Ethiopia; 1,640,000 to 1,970,000 in India; 330,000 to 336,000 in Nigeria; and 111,000 to 133,000 in Pakistan. When harmonising model assumptions, alignment on HPV type distribution significantly narrowed the differences in vaccine impact estimates.

**Conclusions:** The main difference in estimates for cases, deaths, and DALYs averted by vaccination between the models are due to variation in interpretation around data on cervical cancer attribution to HPV-16/18; differences in estimates for DALYs averted are additionally due to differences in age-specific remaining life expectancy over time between the two models. As countries make progress towards the World Health Organization targets for cervical cancer elimination, continued explorations of underlying differences in model inputs, assumptions, and results when examining cervical cancer prevention policy will be critical.

## Background

Persistent infections with human papillomavirus (HPV) types 16 and 18 cause 70% of all cases of cervical cancer [1, 2]. Studies have shown that prophylactic HPV vaccination provides almost 100% protection against persistent infection with vaccine-targeted high-risk HPV strains (e.g., HPV-16, -18) and associated pre-cancers if administered prior to sexual initiation [3, 4]. The World Health Organization (WHO) has set a goal to eliminate cervical cancer as a public health problem by 2100, which involves reducing country-level annual cervical cancer incidence to below 4 per 100,000 [5-7]. However, vaccine coverage remains low in low- and middle-income countries (LMICs) that also lack high quality screening programs [8]. More than 85% of cervical cancer deaths occur in LMICs [9] with cervical cancer being the leading cause of female cancer death in sub-Saharan Africa [10].

Cervical cancer imposes the second greatest burden of cancer incidence among women in Ethiopia, India, Nigeria, and Pakistan, as well as the second greatest burden of cancer mortality among women in Ethiopia, India, and Nigeria—and the fourth greatest burden of cancer mortality among women in Pakistan [11]. The International Agency for Research on Cancer (IARC) estimated that 148,000 new cases occurred and 94,000 women died from cervical cancer in these four countries in 2020 [11]. However, of these four countries, only Ethiopia has implemented nationwide HPV vaccination, with a single-age cohort campaign of 14-year-old girls in 2018–2019 [8, 12], while India has introduced it in a few states in 2016 [13].

Most new vaccine introductions in those countries have been supported through partnerships with the global community, particularly Gavi, the Vaccine Alliance, and the Bill and Melinda Gates Foundation (BMGF). Mathematical models of the impact of HPV vaccines have been used to support programme monitoring and priority-setting by Gavi, the Vaccine Alliance, and BMGF since 2011 [14]. The Vaccine Impact Modelling Consortium (VIMC) was formed in late 2016, with the support of Gavi and BMGF, to bring together two groups involved in HPV modelling, with others conducting impact modelling of 11 other vaccines [15]. Additionally, by ensuring that at least two groups model each analysed pathogen within VIMC, the vaccine impact estimates from VIMC provide an important opportunity to examine the parametric, structural, model, and methodological uncertainty both within and between models [16]. Comparative modelling aims to enhance model transparency and can help guide public health research and priorities.

The two HPV vaccine models in VIMC are the Papillomavirus Rapid Interface for Modelling and Economics (hereafter PRIME), developed by a consortium of modellers led by the London School of Hygiene & Tropical Medicine (LSHTM); and the Harvard model (hereafter Harvard), developed by a team of modellers at the Harvard T.H. Chan School of Public Health. Both models have been used extensively to inform decisions by Gavi [17, 18], BMGF [19], WHO [20-22], and individual countries [23, 24]. Given that both models are being used to understand HPV vaccine impact in the same settings, it is important to quantitatively compare the projections made by both models and to understand their differences, so that model results can be interpreted in the context of each other.

Our study objective was to estimate the potential health impact of HPV vaccination among ten cohorts of 9-year-old girls from 2021–2030 in Ethiopia, India, Nigeria, and Pakistan using two independent mathematical models, and assess similarities and differences in vaccine impact projections through a comparative modelling analysis. We used a vaccination scenario of 90% annual coverage among 9-year-old girls, in alignment with the goals of the cervical cancer elimination strategy set forth by the WHO [5-7]. We estimated the potential health impact in terms of cervical cancer cases, deaths, and disability-adjusted life years (DALYs) averted among vaccinated cohorts from the time of vaccination until 2100 in Ethiopia, India, Nigeria, and Pakistan. We also conducted a comparative modelling analysis to infer the differences in the vaccine impact estimates generated by the PRIME and Harvard models.

## Methods

We used the PRIME and Harvard models to project the impact of HPV vaccination in four high-burden countries (Ethiopia, India, Nigeria, and Pakistan). Both the PRIME [22, 25] and Harvard [17, 23] models have been extensively described and validated elsewhere; we summarize their main features below.

### Model overview

Both the Harvard and PRIME models are static, multi-cohort, proportional impact models that can estimate the impact of HPV vaccination on cervical cancer cases and deaths. The models estimate vaccination impact in terms of reductions in age-dependent cervical cancer incidence and mortality in direct proportion to vaccine efficacy against HPV-16/18, vaccine coverage, and HPV type distribution.

The models assume that girls are fully immunized with a two-dose schedule with perfect timeliness at the target ages and that girls effectively immunized against vaccine-targeted HPV types can develop cervical cancer associated with non-vaccine HPV types; also, no cross-protection against non-vaccine types nor indirect effects are assumed. The model captures burden from all HPV genotypes, but the impact of vaccination is limited to the burden caused by genotypes targeted by the vaccine. In this analysis, the models simulated health benefits from vaccination against HPV types 16 and 18. Vaccine efficacy against HPV-16/18 infections is assumed to be 100% [26-30] over the lifetime. Herd effects are not considered so the vaccine impact estimates produced are conservative. The models assume that age-specific cervical cancer incidence among unvaccinated women remains constant over the time horizon of the model.

### Data sources

Table 1 outlines the data sources used by the Harvard and PRIME models. Age-specific cervical cancer incidence is estimated from the Globocan 2020 database of IARC [11]. For the proportion of cancer that is attributed to the vaccine-covered types (e.g., HPV-16/18), PRIME uses a study by Serrano et al. [31] whose data sources include a meta-analysis performed by the IARC [32] and a retrospective cross-sectional worldwide study [33], while Harvard uses the meta-analysis by the IARC [32] exclusively.

**Table 1.**
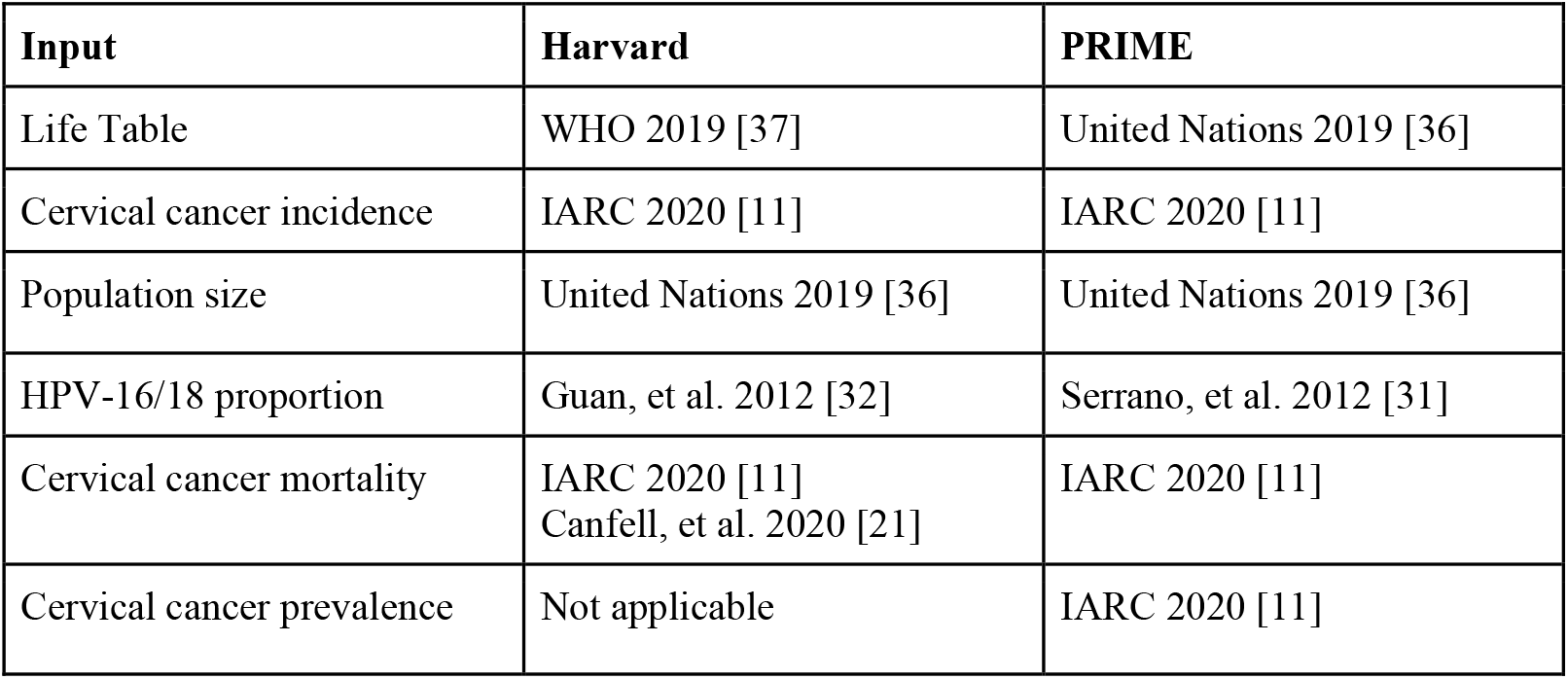
Data sources for Harvard and PRIME models.

To estimate cancer mortality, the Harvard model assumes country-specific distributions of cancer stages [21]. The model then incorporates 5-year stage-specific survival probabilities for untreated and treated cervical cancers (by region) and treatment access proportions (by country). These values are combined into weighted averages to provide country-specific 5-year survival parameters by stage, validated against age-specific mortality rates [11, 21]. The PRIME model uses estimates of age-specific cervical cancer mortality from Globocan 2020 [11].

In the Harvard model, disability weights are assumed to be 0.2 for stages I-III cervical cancer and 0.4733 for stage IV cervical cancer, based on the Global Burden of Disease (GBD) studies [34, 35], and all cervical cancer cases experienced an average of two years lived with disability. In PRIME, disability weights are also based on GBD studies for the different phases of cervical cancer: diagnosis and primary treatment phase (0.288), controlled phase (0.049), metastatic phase (0.451), and terminal phase (0.540). The disability weights and durations for the different phases of cervical cancer are used in estimating the years of life lost due to disability (YLD).

### Demography

PRIME is a multiple cohort model. It calculates population size by estimating the size of the female age cohort at the age of vaccination (e.g., 9 years old) from United Nations Population Division estimates [36]. The size of the age cohort in subsequent years is then calculated by constructing life tables, using the time-varying probability of dying by age and country from the United Nations Population Division estimates [36].

In contrast, the Harvard model is a population-based model that uses only data from the United Nations Population Division [36]. The base year (e.g., 2010) is used for the population projections in year 0, the next year (e.g., 2011) is used for the population projections in year 1 and so forth. Life tables from the World Health Organization are used for calculating DALYs, but not for population projections. Demographic estimates for age-specific population size (in 1-year intervals) and age-specific life expectancy (in five-year intervals) were from United Nations World Population Prospects 2019 revision [36] and 2019 WHO life tables [37], respectively. In years when no data were available, we used a growth factor calculated as a function of a country’s population.

### Vaccination scenarios

We conducted analyses to evaluate the impact of HPV vaccination assuming 90% coverage of annual, routine vaccination of 9-year-old girls vaccinated in 2021–2030 (i.e., ten cohorts). We assumed 100% protection against HPV-16 and -18 infections over the lifetime of vaccinees for a two-dose vaccination schedule.

### Model outcomes

Cervical cancer cases, deaths, and DALYs averted were calculated in comparison with a strategy of no HPV vaccination in the four high-burden countries (Ethiopia, India, Nigeria, and Pakistan) using both PRIME and Harvard models. Model outcomes were aggregated over multiple birth cohorts to capture the health benefits of vaccinating girls aged 9 years between 2021 and 2030 from the time of vaccination until 2100. Model inputs, including population demography and HPV-16/18 type distribution, were harmonised by aligning on the respective input from the respective model in order to evaluate model differences.

## Results

### Descriptive model differences

Figure 1A shows the differences between the data used for the proportion of cervical cancer that is attributed to HPV-16/18 by the two models for Ethiopia, India, Nigeria, and Pakistan. Figure 1B compares the cohort size for a cohort born in 2012 (which relates to the vaccinated cohort of 9-year-old girls in 2021) in the two models using Pakistan as an example; all four countries showed similar differences and trends. While, on average, the differences in cohort size range from 2% to 6%, the differences remain small until older ages (80 years and above), when the differences increased due to the decreasing population size with increasing all-cause mortality.

**Figure 1.**
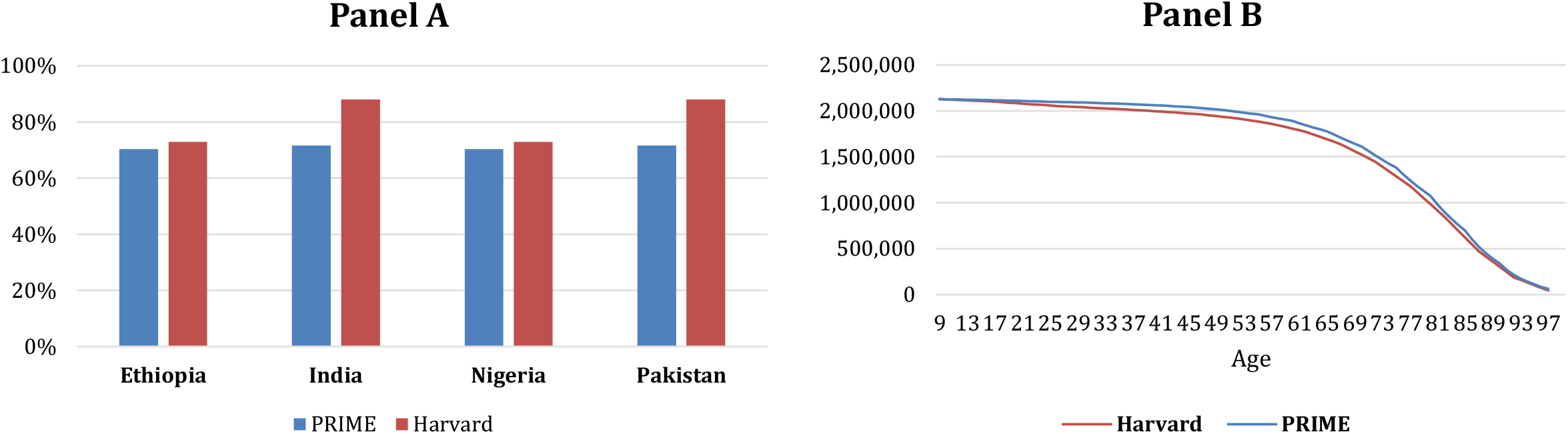
**(A) Proportion of cervical cancer attributable to HPV-16/18; and (B) population size over time for 9-year-old girls born in 2012 (vaccinated in 2021) in Pakistan**.

### Cervical cancer cases, deaths and DALYs averted

Under different assumptions for HPV-16/18 type distribution and demography, the Harvard model estimated a greater number of cervical cancer cases averted than the PRIME model by 3% in Ethiopia, 20% in India, 2% in Nigeria, and 19% in Pakistan (Figure 2A). Specifically, the range between the PRIME model and the Harvard model for the potential health impact of HPV vaccination in terms of the number of cervical cancer cases averted among girls vaccinated 2021– 2030 between the year of vaccination and 2100 was: 262,000 to 270,000 in Ethiopia; 1,640,000 to 1,970,000 in India; 330,000 to 336,000 in Nigeria; and 111,000 to 133,000 in Pakistan.

**Figure 2.**
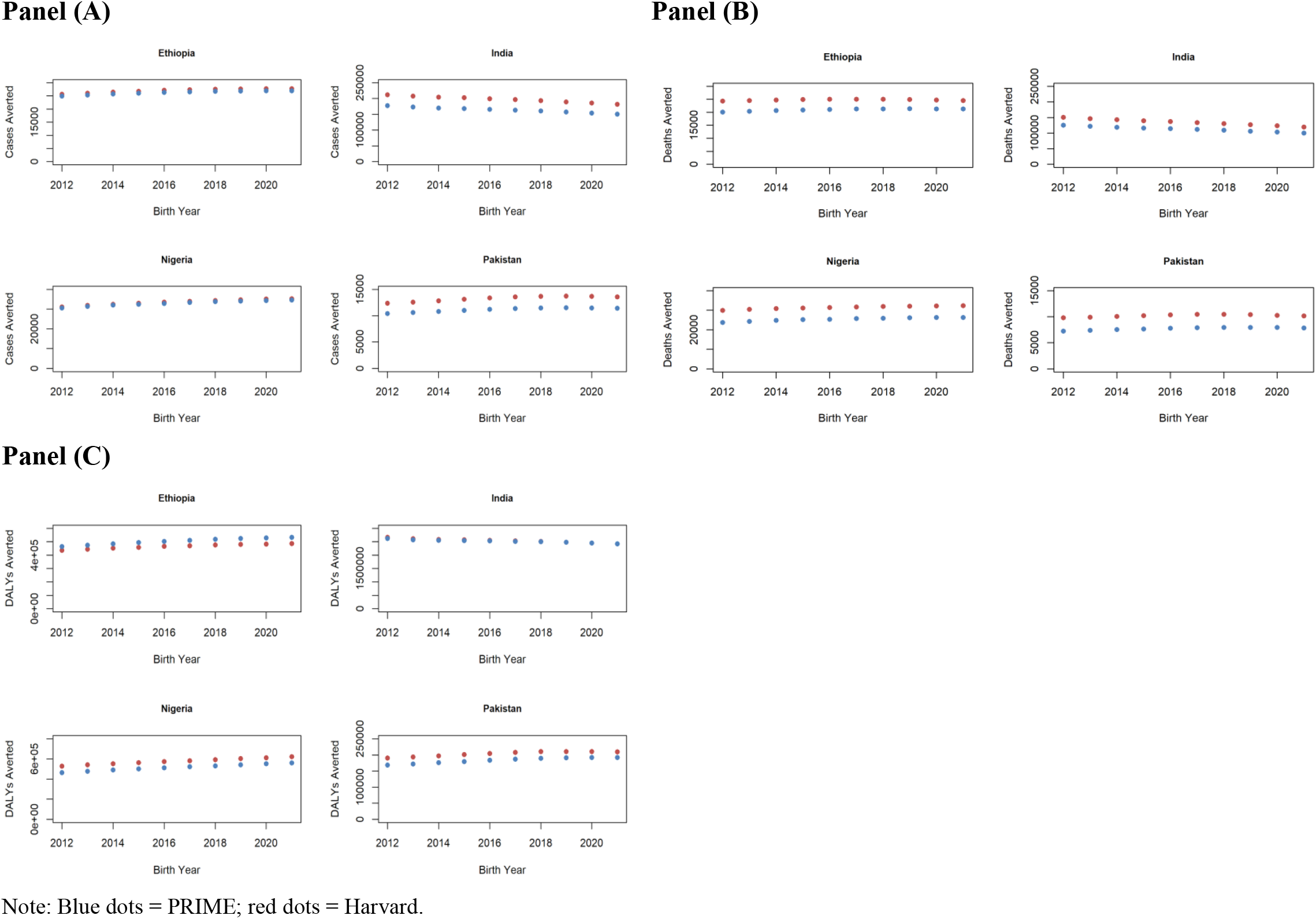
**Cervical cancer cases, deaths, and disability-adjusted life years (DALYs) averted among girls vaccinated during 2021–2030 by country since time of vaccination until 2100: (A) cases averted; (B) deaths averted; (C) DALYs averted**.

Similarly, the Harvard model estimated a greater number of cervical cancer deaths averted than the PRIME model by 15% in Ethiopia, 17% in India, 19% in Nigeria, and 24% in Pakistan (Figure 2B). Specifically, the estimated number of cervical cancer deaths averted ranged from 210,000 to 248,000 in Ethiopia; 1,130,000 to 1,350,000 in India; 254,000 to 314,000 in Nigeria; and 77,200 to 102,000 in Pakistan in the PRIME and Harvard models, respectively.

However, the PRIME model estimated a greater number of DALYs averted than the Harvard model by 8% in Ethiopia; whereas, the Harvard model estimated a greater number of DALYs averted than the PRIME model by 1% in India, 11% in Nigeria, and 10% in Pakistan (Figure 2C). Specifically, the range between the PRIME model and the Harvard model for the estimated number of cervical cancer DALYs averted was: 4,650,000 to 5,030,000 in Ethiopia; 25,200,000 to 25,400,000 in India; 5,160,000 to 5,770,000 in Nigeria; and 1,830,000 to 2,040,000 in Pakistan.

### Vaccination impact

Figure 3 shows the number of cervical cancer cases prevented per 1,000 fully vaccinated girls in each of the four countries compared using both Harvard and PRIME models. When comparing the models under the respective model assumptions for HPV type distribution and population demography, the estimated vaccination impact per 1,000 fully vaccinated girls for the Harvard and PRIME models, respectively, was: 19 vs. 18 in Ethiopia; 20 vs. 16 in India; 12 vs. 12 in Nigeria; and 6 vs. 5 in Pakistan. When harmonising the assumptions around population demography across the Harvard and PRIME models, the difference in the estimated number of cervical cancer cases given routine vaccination narrowed, but the difference in the estimated vaccination impact was slightly increased (Figure 3A). However, overall, the effect of harmonising the assumptions around population demography between Harvard and PRIME was small based on this metric, resulting in an equivalent estimate of the number of cervical cancer cases averted per 1,000 fully vaccinated girls as the base case Harvard and PRIME models. On the other hand, when harmonising the assumptions around HPV-16/18 type distribution between the Harvard and PRIME models (Figure 3B), the differences in estimated vaccination impact were nearly eliminated. Thereby, we infer that the main difference in estimates for cases averted by vaccination between the two models is due to variations in cervical cancer attribution to HPV-16/18.

**Figure 3.**
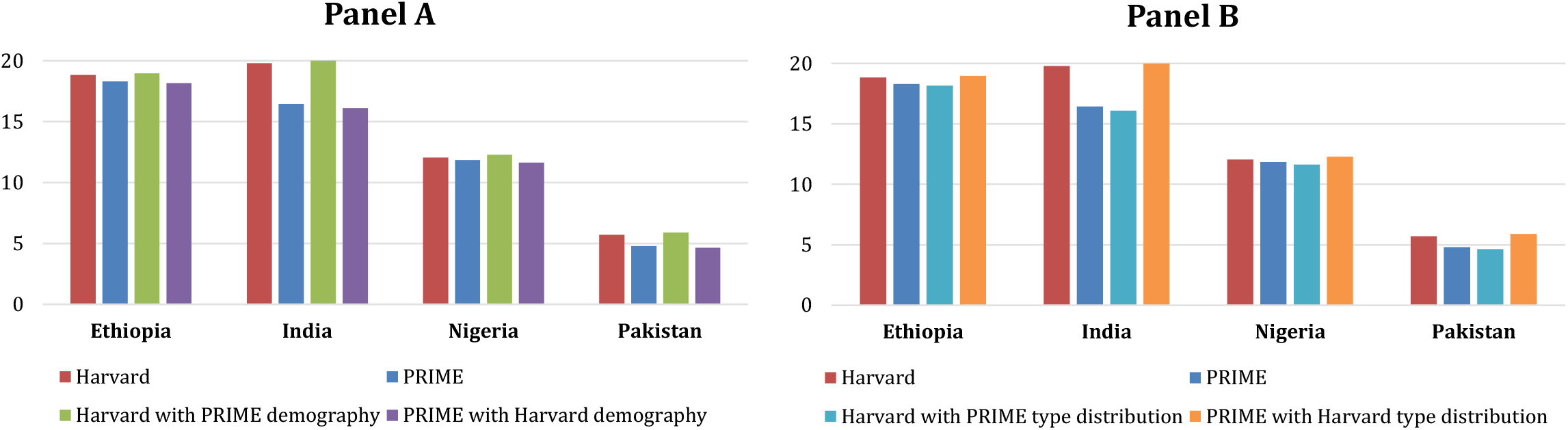
**Cervical cancer cases averted per 1**,**000 fully vaccinated girls for cohorts vaccinated during 2021–2030 since time of vaccination to 2100: (A) with alignment on population demography; and (B) with alignment on HPV 16/18 type distribution**.

## Discussion

The Harvard and PRIME models are used by both VIMC and other global stakeholders to project the impact of HPV vaccination. The two models differ in their inputs and assumptions for HPV-16/18 type distribution, population demography, cervical cancer mortality, and estimation of DALYs. The proportion of cervical cancers due to HPV-16/18 is relatively higher in the Harvard model, especially for India and Pakistan, and thereby cases averted (Figure 2A) and vaccination impact (cases averted per 1,000 fully vaccinated girls) are relatively higher (Figure 3) in the Harvard model for India and Pakistan. The difference between these two models captures variation around interpretation of input data. In the case of HPV-16/18 type distribution, the Harvard model relied on a meta-analysis of cross-sectional high-risk HPV-type distribution in HPV-positive women [32], whereas the PRIME model relied on an estimation of invasive cervical cancer based on this same meta-analysis [32] that accounts for multi-type infections through proportional weighting attribution [31].

HPV-16/18 vaccination was estimated to avert substantial numbers of cervical cancer cases, deaths, and DALYs by both the Harvard model and the PRIME model. However, routine HPV vaccination has yet to be introduced in any of these four high-burden countries at the national level (although a nationwide single-age cohort campaign of 14-year-old girls was conducted in Ethiopia in 2018–2019 [12] and India introduced in a few states in 2016 [13]). In order to accelerate progress towards cervical cancer elimination, preventing cervical cancer through HPV vaccination will be an essential strategy in these countries, particularly given the low coverage and access to cervical cancer screening [8]. Understanding similarities and differences between HPV vaccination impact predicted by different models will be crucial, given that key questions about which countries to prioritise and which vaccination strategies to use will be important in an era of HPV vaccine dose shortages [38] and COVID-19-related disruptions to vaccination programmes [39, 40].

The size of the relevant population at each year of age is critical because it determines the number of people who are exposed to the risk of cervical cancer and can therefore be protected by vaccination. Both methods used to estimate the at-risk population have strengths and limitations. In the PRIME model, the size of the age cohort in subsequent years is calculated by constructing life tables, using the time-varying probability of dying by age and country from the United Nations Population Division estimates [36]. The Harvard method captures such changes, but also includes future population change due to migration, which may not reflect the same vaccination status as the locally-born population. In general, as the demography differences are minimal between the two models, the vaccination impact estimates are similar for each country when demography is switched to align with the alternative model. The main driver of the differences in the vaccination impact was HPV-16/18 type distribution—or, the proportion of cervical cancers that can be averted by the bivalent vaccine, which has a direct relationship with vaccination impact.

This comparative modelling exercise highlighted differences in the estimates of health impact of HPV vaccination due to model uncertainty. We note that HPV vaccine projections are particularly susceptible to large swings in estimated health outcomes due to even small changes in fertility and mortality because of the long time horizons needed in the models. Comparative modelling exercises as we have done can enhance model transparency and clarify the range of uncertainty in vaccine impact. Hence, the differences between the models are a strength that reflects the variation and uncertainty in the projected health outcomes of vaccination impact. Understanding the inter-model variation improves the quality and coordination of vaccine impact assessment, which in turn can help guide public health research and priorities in cervical cancer elimination and control.

Similar comparative modelling exercises were conducted to examine the timeline to cervical cancer elimination in LMICs [20, 21]. Relying on evidence synthesis from different models was deemed an essential aspect to inform strategies for cervical cancer elimination by the WHO [5, 41]. However, these analyses relied on estimations of age-standardized cervical cancer incidence, such that demographic changes were not expected to drive the uncertainty in the timing of elimination.

There are several important limitations to this analysis. As we relied on static cohort models in these analyses, we were only able to estimate direct effects for vaccinated women, which excluded additional indirect benefits from herd immunity for unvaccinated women. We projected intervention impact for only ten cohorts of 9-year-old girls in four countries and assumed that cervical cancer incidence rates affecting these cohorts would be stable over the time period of the analysis. Vaccine efficacy against high-risk HPV types other than HPV-16/18 (i.e., cross-protection) was not included. We did not examine cervical cancer screening programs in this analysis and assumed that any ongoing screening programs did not change as HPV vaccination introduction and delivery changed. Additionally, given limited data on the burden of other HPV-related diseases in LMICs, we did not evaluate the impact HPV vaccination may have on non-cervical cancers in women and men.

## Conclusions

Both models project that HPV vaccination will have a large impact on morbidity and mortality in the four countries we examined. The differences in outcomes between the models capture variation in interpretation around data on cervical cancer epidemiology and future demographic change. This study highlights that HPV type distribution is a critical input to modelling the potential health impact of vaccination. The main difference in estimates for cases and deaths averted by vaccination between the models capture variation in interpretation around data on cervical cancer attribution to HPV-16/18. The main differences in estimates for DALYs averted by vaccination between the models are due to variations in cervical cancer attribution to HPV-16/18 and age-specific remaining life expectancy over time. Continued explorations of underlying differences in model inputs, assumptions, and results will be crucial when examining public health policy.

## Data Availability

The datasets generated during and/or analysed during the current study are available from the corresponding author on reasonable request.

## List of abbreviations

BMGF: Bill & Melinda Gates Foundation
DALY: disability-adjusted life year
GBD: Global Burden of Disease
HPV: human papillomavirus
IARC: International Agency for Research on Cancer
LMICs: low- and middle-income countries
LSHTM: London School of Hygiene & Tropical Medicine
PRIME: Papillomavirus Rapid Interface for Modelling and Economics
VIMC: Vaccine Impact Modelling Consortium
WHO: World Health Organization
YLD: years of life lost due to disability

## Declarations

### Ethics approval and consent to participate

Not applicable.

### Consent for publication

Not applicable.

## Authors’ contributions

AP, KA, and MJ conceptualized and designed the study. AP and KA did the analysis, drafted the initial manuscript, and approved the final manuscript as submitted. SS, JJK, and MJ critically reviewed the analysis, reviewed and revised the manuscript, and approved the final manuscript as submitted.

## Acknowledgements

We would like to thank Caroline L. Trotter and Alicia N.M. Kraay who provided valuable comments during a review of these models organized by the Vaccine Impact Modelling Consortium.

